# COVID-19 spreading in Rio de Janeiro, Brazil: do the policies of social isolation really work?

**DOI:** 10.1101/2020.04.27.20081737

**Authors:** Nuno Crokidakis

## Abstract

The recent Coronavirus (COVID-19) has been spreading through all the world fastly. In this work we focus on the evolution of the COVID-19 in one of the most populous Brazilian states, namely the Rio de Janeiro state. The first case was reported in March 5, 2020, thus we have a considerable amount of available data to make a good analysis. First we study the early evolution of the disease, considering a Susceptible-Infectious-Quarantined-Recovered (SIQR) model. This initial phase shows the usual exponential growth of the number of confirmed cases. In this case, we estimate the parameters of the model based on the data, as well as the epidemic doubling time. After, we analyze all the available data, from March 5, 2020 through April 26, 2020. In this case, we observe a distinct behavior: a sub-exponential growth. In order to capture this change in the behavior of the evolution of the confirmed cases, we consider the implementation of isolation policies. The modified model agrees well with data. Finally, we consider the relaxation of such policies, and discuss about the ideal period of time to release people to return to their activities.

## I. INTRODUCTION

The world observed recently the emergence of a new pandemic, the COVID-19 (coronavirus 2) caused by severe acute respiratory syndrome SARS-CoV-2 [1]. As other pandemics observed during the time, COVID-19 has been caused a lot of fatal victims. The emergence of diverse epidemics led the scientific community to developed diverse methods to study and analyze the evolution of diseases. The classical compartmentals mathematical models are usually the starting point for the study of epidemics [2, 3].

In the last two months a considerable number of works and preprints were published considering studies related to the COVID-19 evolution through the world [4–30]. Some of such papers considered the case of Brazil. Our interest in this work is to study the evolution of COVID-19 in one of the most populous Brazilian states, namely the Rio de Janeiro state. We collected data from the Brazilian Department of Health, and apply a compartmental model to analyze the COVID-19 dynamics.

The first case in the Rio de Janeiro state was reported in the Rio de Janeiro city in March 5, 2020. After that, the disease spreaded to all the State, achieving the majority of state’s cities. To avoid the rapid growth of the number of cases, the Rio de Janeiro state government implemented some policies of social isolation. Such policies were started at March 17, 2020, and of course they lead some days to produce effective results. The government did not imposed a lockdown, they suggested people to stay in home, schools and universities were closed, public events were banning, and only the essential activities are in progress, for example delivery of food, drugstories, supermarkets, and so on.

In order to make an analysis of such isolation policies, we consider a Susceptible-Infectious-Quarantined-Recovered (SIQR) model, and in addition we consider the implementation of isolation in a simple way. The modified model can describe well the sub-exponential growth of the number of cases exhibited by the data. We also discuss about the relaxation of the isolation policies, since that is in discussion by the Rio de Janeiro state government.

This work is organized as follows. Section II is divided in two subsctions. In the first one, we present the SIQR model, and define its parameters, in order to study the early evolution of the COVID-19 in Rio de Janeiro state. In the second subsection we consider the implementation of isolation policies, and study their impact on the dynamics of the SIQR model. Finally, we present a discussion in Section III. Some other details are presented in the Appendix.

## II. MODELS

### A. Early evolution of COVID-19

First, we wil consider the early evolution of COVID-19 in Rio de Janeiro state. As pointed in the Introduction, the first documented case occurred in March 5, 2020 [31]. So, we considered the first days of the evolution of the disease, namely from March 5, 2020 through March 24, 2020. As it is usual, we expect an exponential growth of the number of confirmed cases in such initial phase of the epidemics.

To study the initial evolution of the COVID-19, we consider a Susceptible-Infectious-Quarantined-Recovered (SIQR) model [6, 7, 24, 32]. The total population is *N*, and we consider it constant since we are in the early phase of the epidemic (*N* = *S* + *I* + *Q* + *R*).

As discussed previously [6], both the Infected (I) and Quarantined (Q) individuals are infected. The separation of those individuals in two classes is convenient for the modeling of COVID-19, since it has been reported that there are many individuals that does not developed symptoms of the disease [4]. In addition, the governments (including the Rio de Janeiro governor’s) are forcing individuals tested positive (confirmed cases) to self-isolate from the community (quarantine). In this case, the SIQR model may be described by the following equations:

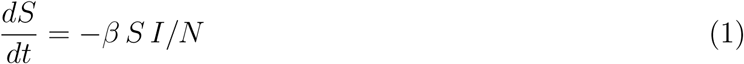

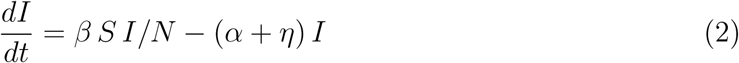

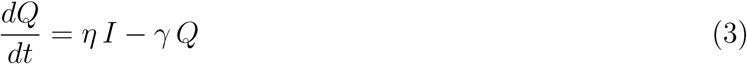

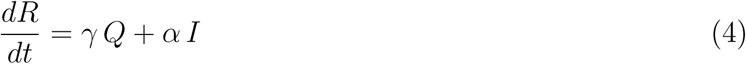

Eqs. (1) - (4) describes the dynamics of the disease spreading, taking into account the parameters *β*, *α, η* and *γ*, denoting the infection rate, the recovering of asymptomatic individuals, detection of infected individuals and recovering of quarantined individuals, respectively.

Following the analytical considerations of Ref. [24], one can derive the expression

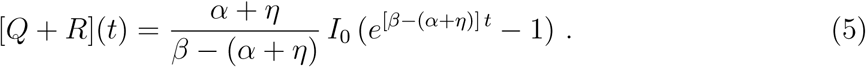

We fitted Eq. (5) to the the Rio de Janeiro state COVID-19 data [31] from March 5, 2020 through March 24, 2020. The estimated values were (*α* + *η*) *I*_0_ = 0.3303 and *β* − (*α* + *η*) = 0.2782 (for details of the fitting procedure, see [24]). First, we can estimate *I*_0_, the number of infectious individuals at the beggining of the outbreak. Based on the data, we take *I*_0_ = 13. Considering this value, the above fitted results give us *α* + *η* = 0.0254 and *β* = 0.3036. Following the discussion in our previous work [24], we have *η* = *α* = 0.0127 and *γ* = 0.02. Considering those estimates, we plot in Fig. 1 the temporal evolution of the number of cases together with Eq. (5).

**FIG. 1.**
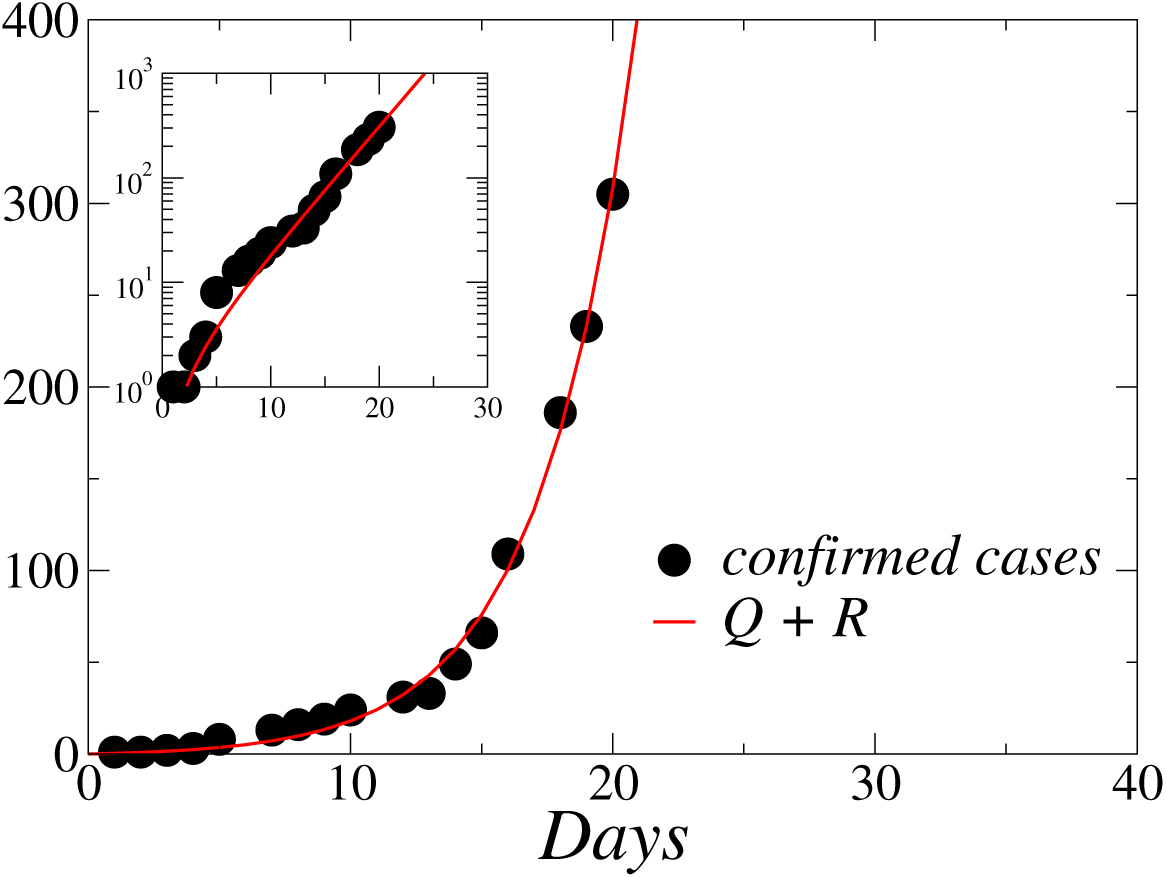
(Color online) Comparison between the number of confirmed cases in Rio de Janeiro state (circles) and Eq. (5) (full line), for the initial epidemic phase. The parameters are *I*_0_ = 13, *β* = 0. 3036, *α* + *η* = 0.0254, as discussed in the text. In the inset we exhibit the graphic in the log-linear scale, showing the typical linear behavior of the exponential function.

One can also estimate the epidemic doubling time, that characterize the sequence of intervals at which the cumulative incidence doubles. Following [24], the doubling time is given by

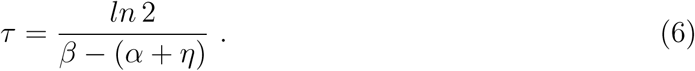

Based on Eq. (6), and on the estimated parameters, we have *τ* ≈ 2.5 days, which falls in the range 1.4 < *τ* < 3.0 estimated in China [18] and agrees with the estimate for Brazil, namely *τ* = 2.72 [24].

Considering the estimated parameters, we exhibit in Fig. 2 the time evolution of the number of Infected (I), Quarantined (Q) and total confirmed cases (Q+R), obtained by the numerical integration of the Eqs. (1) to (4). For these curves, we considered *N* as the total population of the Rio de Janeiro state, *N* = 1.646 × 10^7^. One can see that the number of infected and nonconfirmed cases I grows faster than the number of quaratined individuals Q (confirmed cases). This unbalance is observed in all the world, since there is a huge number of undocumented infection cases for the COVID-19, as discussed in a recent work [4].

**FIG. 2.**
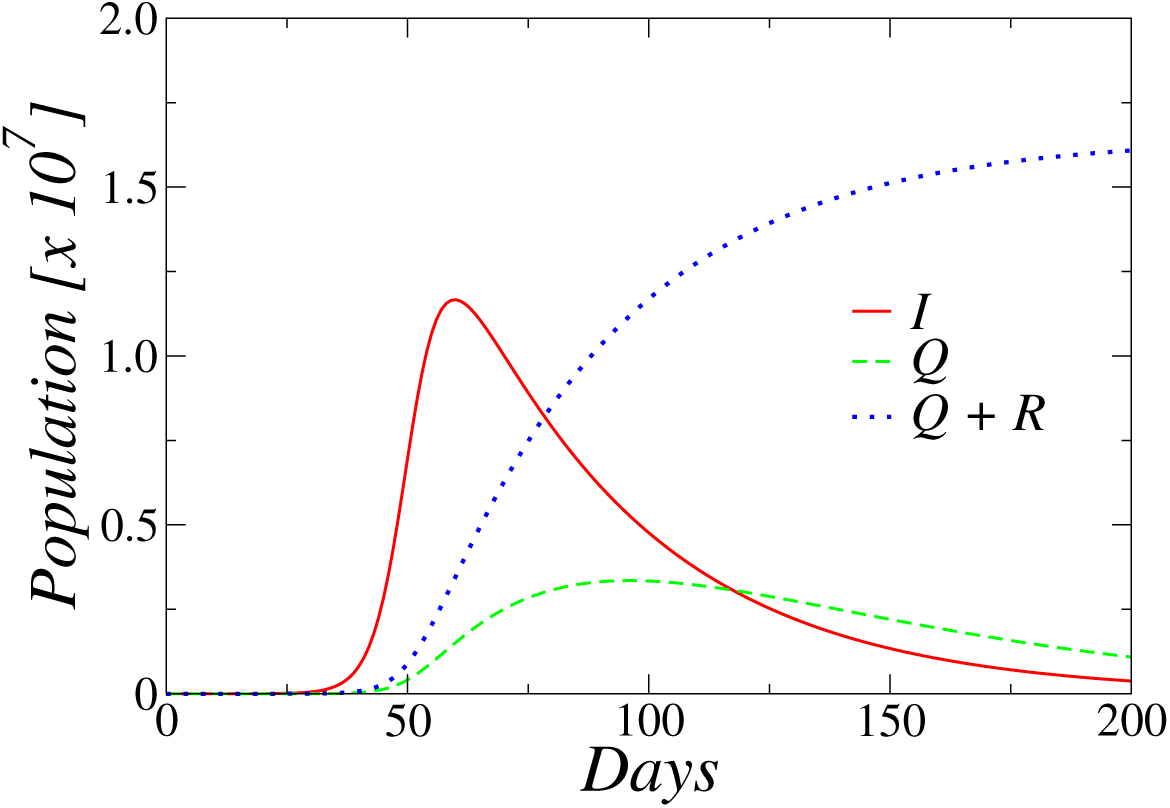
(Color online) Time evolution of the number of Infected (I), Quarantined (Q) and total confirmed cases (Q+R) predicted by the SIQR model, for the initial epidemic phase, obtained by the numerical integration of Eqs. (1) to (4). The parameters are *I*_0_ = 13, *β* = 0.3036, *η* = *α* = 0.0127 and *γ* = 0.02.

The above-mentioned unbalance among I and Q individuals can be estimated through the relation [7, 24]

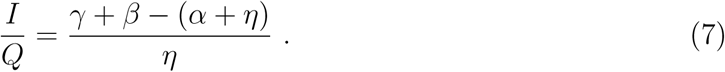

Eq. (7) gives us *I*/*Q* ≈ 23.5, i.e., for each patient in quarantine approximately 23 infectious individuals are present in the population. This result is in good agreement with results reported for countries like China [4] and Brazil [33].

### B. Recent evolution of COVID-19

In Fig. 3 (left side) we plot the data of confirmed cases of COVID-19 for the Rio de Janeiro state, now considering all the evolution of cases, from March 5, 2020 through April 26, 2020, obtained from [31]. For comparison, we also plot Eq. (5) (dasehd line) together with data of the initial epidemic phase. As one can see, the behavior is not purely exponential, as was the case for the early growth of cases discussed in the previous subsection. This is consequence of the policies of social isolation implemented by the Rio de Janeiro state governement. Indeed, the number of confirmed cases is still growing, but this growth is slower than the initial exponencial behavior. We observed a linear behavior when we plot the data in the log-log scale, as observed in the data from several provinces of China [29], as well for some other countries [34]. Thus, to guide the eyes, we also exhibit a power law function *f*(*t*) = *ct^d^* (full line)^1^. As one can see in Fig. 3 (left side), the confirmed cases are growing sub-exponentially. We also exhibit in Fig. 3 (right side) the same curves in the log-linear scale. The vertical dotted line marks the beginning of the isolation policies in Rio de Janeiro (March 17, 2020). One can see that the social distancing policies changed the initial exponential growth after 7 days (March 24, 2020).

**FIG. 3.**
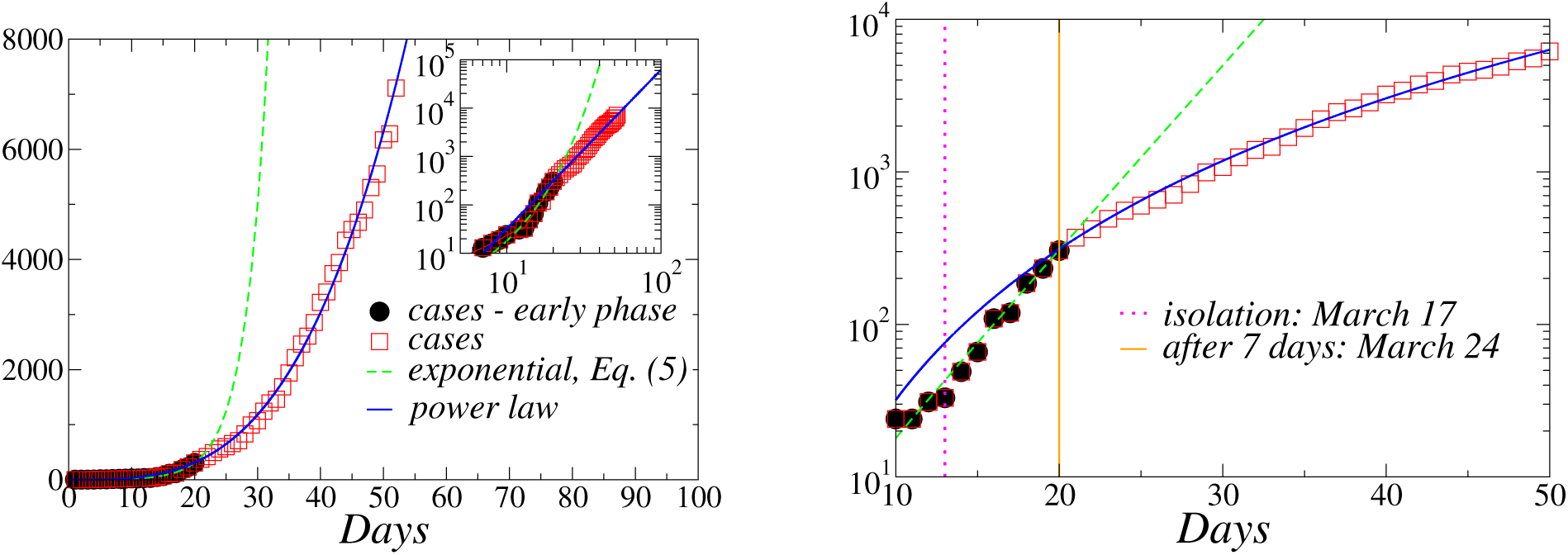
(Color online) *Left panel*:Time evolution of the total number of confirmed cases from March 5, 2020 through April 26, 2020 (squares). The dashed line is given by Eq. (5). As a guide to the eyes, we also show a power law fit of the data (full line). In the inset we exhibit the same data in the log-log scale. *Right panel*: Log-linear plot of the same quantities. The vertical dotted line marks the beginning of the isolation policies in Rio de Janeiro (March 17, 2020). One can see that the social distancing policies changed the initial exponential growth after 7 days (March 24, 2020).

In this case, to reproduce the mentioned sub-exponential behavior of the number of confirmed cases, we have to modify the SIQR model. As previous discussed, the sub-exponential behavior may be caused by isolation policies. A simple form to modeling the isolation policies is to consider an aditional mechanism that can be interpreted as a process of removing susceptibles from the transmission process, as discussed recently [29]. Thus, following [29], one can consider a containment rate *k*_0_, such that Eqs. (1) - (4) are modified to

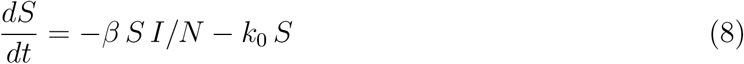

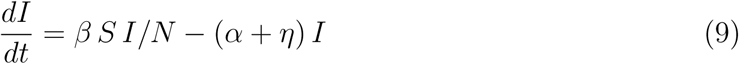

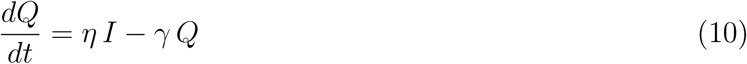

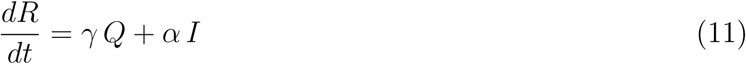

The case *k*_0_ = 0 corresponds the SIQR model discussed in the previous subsection, which represents a scenario in which the general population is unaffected by policies or does not commit behavioral changes in response to an epidemic. This simple mechanism can reproduce the sub-exponential behavior observed in the data, as we will see in the following.

Some insights can be obtained from analytical considerations. Following the equations derived in the Appendix, one can obtain

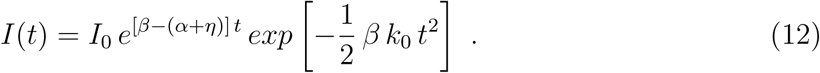

The first factor implies that quarantining infecteds merely decreases the rate with which their number will grow exponentially, as occurred in the SIQR model studied in the previous subsection. On the other hand, the second term suppresses the whole transmission process well enough to alter the growth behavior, due to the quadratic term *t*^2^. This implies that public shutdown policies facilitate epidemic containment in a more effective way than quarantine measures [29].

As we mentioned in the beginning of this subsection, we are not anymore in the early evoluton of COVID-19. In such case, Eq. (12) is a poor approximation for the real evolution of Eq. (9), and as a consequence, a poor approximation for the evolution of the four compartments. Thus, despite one can derive another analytical expression for the total of confirmed cases *Q* + *R*, the estimated parameters will not be accurate as in the previous section, but they can be usefull (see the Appendix).

In Fig. 4 (left side) we plot the total number of confirmed cases of COVID-19 in Rio de Janeiro state, as well as the temporal evolution of the quantity *Q* + *R* predicted by the model. There is a good agreement between data and model for the adjusted parameters *I*_0_ = 24, *β* = 0.32, *η* = *α* = 0.018, *k*_0_ = 0.033 and *γ* = 0.02. For comparison with the model with no isolation policies, we also plot in Fig. 4 (right side) the evolution of the Quarantined (Q) individuals for the above-mentioned parameters, and the same quantity obtained in the subsection II.A. One can see the flattening of the curve: the maximum number of quarantined individuals decreases considerably, even for a small value of *k*_0_.

**FIG. 4.**
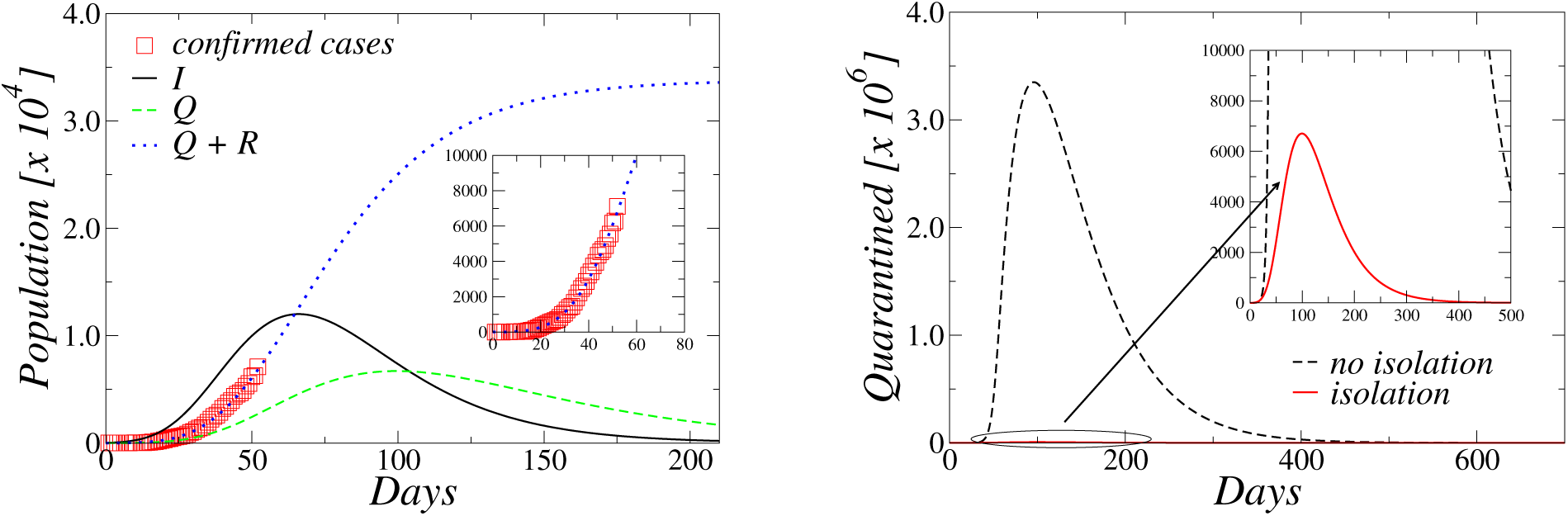
(Color online) *Left panel*: Time evolution of the number of Infected (I), Quarantined (Q) and total confirmed cases (Q+R) considering all the data and isolation policies, obtained by the numerical integration of Eqs. (8) to (11). Data of confirmed cases are exhibited as well (squares). In the inset we show a zoom in the region of the current data (from March 5, 2020 through April 26, 2020), to highlight the agreement of the model with the data. *Right panel*: Flattening the curve: comparison between the predicted number of Quarantined individuals for the model with (full line) and without (dashed line) the implementation of isolation policies. The peak of the curve is considerably reduced. In both figures the parameters are *I*_0_ = 24, *β* = 0.32, *η* = *α* = 0.018, *k*_0_ = 0.033 and *γ* = 0.02.

Considering the result of Fig. 4 (right side), the peak of the Quarantined individuals is expected to occur in the Day 99, i.e., June 11. Some local governments in Brazil are discussing about the relaxation of isolation of people including the Rio de Janeiro state government. A question of practical interest arises: when is it really safe to relax the isolation policies? To address this issue, we considered the following slight modification of the model presented in this section. In this case, the dynamics is implemented normally considering Eqs. (8) - (11) from the initial time *t* = 0 until a given time step *t′*. After *t′*, we take *k*_0_ = 0 in order to simulate the relaxation of isolation policies, and the model is reduced to the SIQR model presented in section II.A. The equations are integrated numerically, and the results for distinct times *t′* are exhibited in Fig. 5. One can see that if the relaxation of isolation policies is started at *t*′ = 61 (May 4, 2020), the result is disastrous: the peak of Quarantined individuals increases fast. However, one can also see in Fig. 5 that after June 1, 2020, there is no considerably change in the peak, suggesting a security date to relax the isolation policies. Even after May 20 the peak does not increase considerably, which lead us to conclude that if the end of isolation procedures occur between May 20 and June 1 the damage will not be considerable.

**FIG. 5.**
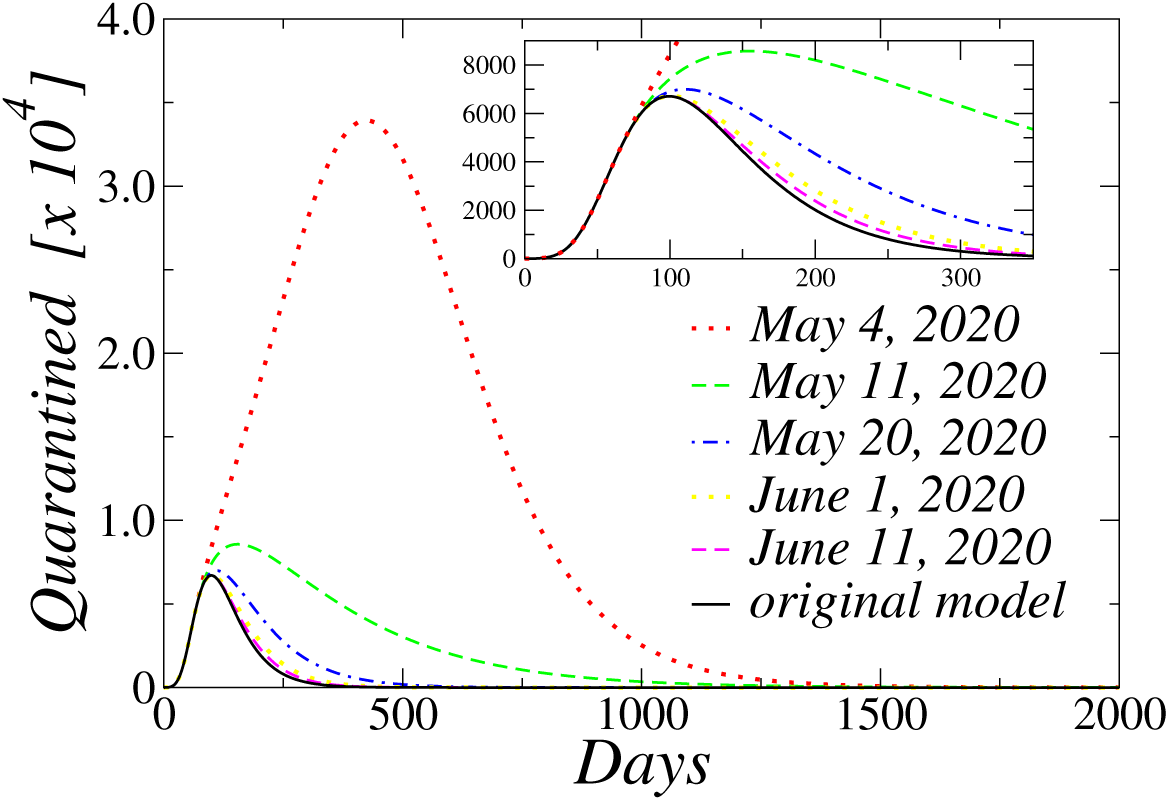
(Color online) Time evolution of the number of Quarantined (Q) individuals. The full line represents the main result of section II.B, i.e., the original model with isolation policies *(k*_0_ = 0.033). The other curves represent the relaxation of such policies, starting from the days *t′* indicated in the graph. The inset shows a zoom of the smallest peaks. One can see that after June 1, 2020 there is no considerably change in the peak.

## III. DISCUSSION

The number of confirmed cases of COVID-19 in the Rio de Janeiro state, Brazil, are still growing. Based on the data, we considered a SIQR on a fully-connected population in order to study the early evolution of the disease. The initial growth of confirmed cases is exponential, as it is usual, and the parameters of such early epidemic phase were estimated, based on the data.

However, as discussed in the text, the Rio de Janeiro state governement implemented isolation policies in March 17, in order to avoid the explosion of cases of COVID-19. The data after the early evolution of COVID-19 exhibits a distinct, nonexponential behavior, i.e., the growth of the number of cases becomes sub-exponential, and the SIQR model can not capture such behavior. In this case, we modified the model to consider isolation procedures in a simple way. The modified model describes well the data, and it allowed us to make a detailed analysis of the evolution of COVID-19 in Rio de Janeiro. Considering the data, we have observed that the initial exponential behavior changed to the sub-exponential one about 7 days after the implementation of social distancing policies.

We found that the isolation policies really work, i.e., the fact of isolate people in home, to avoid social contacts, decreased the growth rate of the number of cases. For the considered parameters, we observed that the number of quarantined individuals grows, stabilizes and after it decays to zero, as it is standard in compartmental models. Based on the data, we can see that the number of such isolated individuals grows until Day 99 from the beginning (March 5, 2020) of the disesase spreading. Thus, the model predicts that the maximum number of quarantined individuals will occur about June 11, 2020. This is in line with a recent estimate [14]. That peak is associated with the isolation of about 0.041% of the Rio de Janeiro state population (about 6, 700 individuals). This number is about 500 times lower than the one predicted by the SIQR model without isolation policies, suggesting the effectiveness of the social isolation. In fact, combining quarantine of individuals tested positive (confirmed cases) with social distance, i.e., multiple interventions, appears to be effective to contain the rapid evolution of confirmed cases seen in the early evolution of the COVID-19 in Rio de Janeiro state. This is in line with a recent work [14], where the authors analyzed the potential role of nonpharmaceutical interventions in UK and USA. They conclude that the effectiveness of any one intervention in isolation is likely to be limited, requiring multiple interventions to be combined to have a substantial impact on transmission.

However, this isolation can not lead an infinite time, due to economic consequences for the state. In this case, the government is discussing about the relaxation of the isolation policies. In this case, we modified the model to consider isolation until a given number of days, namely until a time step *t*′. Thus, after such time step *t*′ the isolation parameter *k*_0_ is taken to zero, simulating the relaxation of people isolation at home. Analyzing distinct values of *t*′, we concluded that if the end of isolation procedures occur between May 20 and June 1 the damage will not be considerably. A recent work, considering Brazil, also suggested that the the social distancing policies need to continued for some time in order to really produce effective results [25].

## Data Availability

All data are available in the link provided in the specific location, as well as in the references of the manuscript.

http://painel.saude.rj.gov.br/monitoramento/covid19.html

## Appendix

In this appendix we derive some approximated analytical results of the model presented in section II.B. Typically in outbreaks a small number of people are infected initially. In this case, we have for small times *S*/*N* ≈ 1 and I ≈ 0, such that Eq. (8) can be linearized, yielding the solution [29]

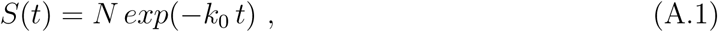

which implies that the depletion of susceptibles available to the transmission process will be dominated by shutdown policies. Considering this result, one can integrate Eq. (9) to obtain

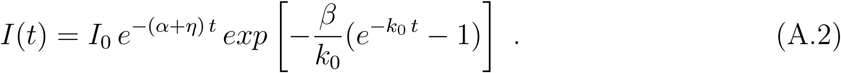

For small values of *t* we can expand the exponential 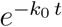 in the last term to obtain the approximate growth function

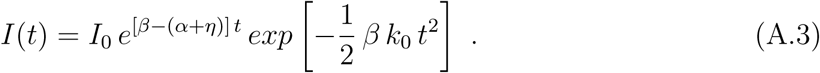

Adding Eqs. (10) and (11), one obtains *d*[(*Q* + *R*)(*t*)]/*dt* = (*α* + *η*) *I*(*t*). Using Eq. (A.3) in this last result and integrating over *t*, one obtains

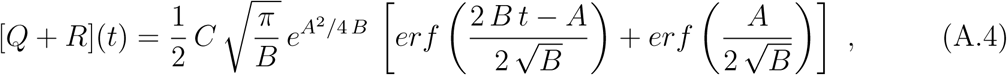

where *er f*(*x*) is the error function, and we have *A* = *β* − (*α* + *η*) and *B* = (1/2) *β k*_0_ and *C* = (*α* + *η*) *I*_0_.

As we mentioned in the text, we are not anymore in the early evoluton of COVID-19. In such case, Eqs. (A.1) - (A.4) are poor approximations for the real evolution of Eqs. (8) - (11). Thus, a fitting procedure based on the data together with Eq. (A.4) will not give accurate parameters, but they can be usefull. In such case, fitting Eq. (A.4) to the data of confirmed cases from March 5, 2020 through April 26, 2020, we obtained *A* = 0.26667 ± 0.01218, *B* = 0.00297 ± 0.00019 and *C* = 0.68921 ± 0.12740. Based on such values, the estimated parameters are *β* = 0.29539, *α* + *η* = 0.02871 (considering *I*_0_ = 24, see the next paragraph) and *k*_0_ = 0.02011. Eq. (A.4) can fit well the data, however if we consider the estimated parameters and perform a numerical integration of the model’s equations (8) - (11), there is a good agreement only for short times. This explains why we fitted the data and analyzed the model in section II.B with other values of the parameters, namely *β* = 0.32, *η* = *α* = 0.018, *k*_0_ = 0.033 and *γ* = 0.02.

Finally, the time in our graphics were counted after the first confirmed case in Rio de Janeiro state, i.e., Day 1 was March 5, 2020. The number of confirmed cases kept almost constant for some days, and start to grow faster since day 10 (March 14, 2020), where 24 cases were confirmed. In this case, we take the number of initial cases as *I*_0_ = 24 for the model’s purposes.

## ACKNOWLEDGMENTS

The author acknowledges financial support from the Brazilian scientific funding agencies Conselho Nacional de Desenvolvimento Científico e Tecnológico (CNPq) and Fundação de Amparo à Pesquisa do Estado do Rio de Janeiro (FAPERJ). Further discussions with Lucas Sigaud are also acknowledged.

## Footnotes

1 The fitting was done just do guide the eyes. We obtained from a least square fitting *c* = 0.0167 ± 0.0031 and *d* = 3.28 ± 0.05.

